# A machine learning approach for precision diagnosis of juvenile-onset SLE

**DOI:** 10.1101/19007765

**Authors:** George A Robinson, Junjie Peng, Pierre Dönnes, Leda Coelewij, Anna Radziszewska, Chris Wincup, Hannah Peckham, David A Isenberg, Yiannis Ioannou, Coziana Ciurtin, Ines Pineda-Torra, Elizabeth C Jury

**Author notes:** Corresponding authors: Liz Jury, Coziana Ciurtin, Ines Pineda-Torra, George Robinson. Share first authorship (George A Robinson, Junjie Peng). Share senior authorship (Coziana Ciurtin, Ines Pineda-Torra, Elizabeth C Jury).

## Abstract

Juvenile-Onset systemic lupus erythematosus (JSLE) is an autoimmune rheumatic disease characterised by systemic inflammation and organ damage, with disease onset often coinciding with puberty. JSLE is associated with more severe disease manifestations and a higher motility rate compared to adult SLE. Due to the heterogeneous clinical and immunological manifestations of JSLE, delayed diagnosis and poor treatment efficacy are major barriers for improving patient outcome. In order to define a unique immunophenotyping profile distinguishing JSLE patients from age matched healthy controls, immune-based machine learning (ML) approaches were applied. Balanced random forest analysis discriminated JSLE patients from healthy controls with an overall 91% prediction accuracy. The top-ranked immunological features were selected from the optimal ML model and were validated by partial least squares discriminant analysis and logistic regression analysis. Patients could be clustered into four distinct groups based on the top hits from the ML model, providing an opportunity for tailored therapy. Moreover, complex correlations between the JSLE immune profile and clinical features of disease were identified. Further immunological association studies are essential for developing data-driven personalised medicine approaches to aid diagnosis of JSLE for targeted therapy and improved patient outcomes.

## Introduction

Systemic lupus erythematosus (SLE) is a chronic, multi-system autoimmune rheumatic disease with a complex aetiology [1]. Juvenile-onset SLE (JSLE) accounts for approximately 15% to 20% of all cases and is defined as the development of SLE in childhood or adolescence [2]. JSLE has a more aggressive disease presentation characterised by increased prevalence of malar rash, nephropathy, central nervous system (CNS) disease and haematological manifestations [3-6] compared to adults with SLE. Severe disease complications, including multiple-organ impairment and a notable increase in risk of cardiovascular disease, are also described in JSLE [7]. To date, no medications have been approved for the treatment of JSLE due to the significant paucity of adolescent-specific trial data [8, 9]. Primarily, the same pharmaceutical drugs are used in JSLE as in adult SLE; however, JSLE patients commonly require increased corticosteroids and immunosuppressive therapies because of their more severe disease presentation [2, 5, 10, 11].

The heterogeneous nature of the clinical manifestations are matched by the broad variety of genetic and immunological abnormalities in JSLE patients [2], making precision diagnosis particularly difficult in these patients. The predicament with imprecise diagnosis and poor treatment efficacy leading to unsatisfactory outcomes for JSLE patients emphasises the urgent need for a widely accepted universal diagnostic criteria and valid patient stratification for personalised treatment.

Recently, in depth computational analysis of large multi-omic datasets has accelerated understanding of complex heterogeneous diseases such as SLE/JSLE [12-15]. Machine learning (ML) is a subdivision of artificial intelligence that builds analytical models by learning by example and has been used in a wide range of clinical areas, including medical image classification and prediction [16], drug discovery by predicting the optimal pharmaceutical target [17], and building predictive models for disease diagnosis and prognosis [18]. It relies upon data collection, data preparation, model training, model evaluation and improved performance cycles for self-improvement, giving it enormous predictive power. A random forest is an ensemble ML algorithm for classification consisting of multitudinous decisions trees, as in a real forest. Compared to a single decision tree, the random forest method can increase the accuracy of the model without suffering from overfitting data [19]. Each decision tree is able to generate independent predictions and vote for the decisive classification outcome, which is more accurate than performing any individual decision tree model alone.

The most recent SLE study applied three different machine learning approaches, including k-nearest neighbours (KNN), generalized logistic models (GLM) and random forest models to predict disease activity using gene expression profiles. The random forest classifier outmatched other approaches by achieving an 83% accuracy under 10-fold cross-validation [20]. Another study used random forest models to predict lupus nephritis outcomes [21].

Here, ML approaches were applied to immunophenotyping profiles of JSLE patients and age-matched healthy controls. An immunological signature was identified and validated using a predictive ML model. This signature could be translated into potential diagnostic and therapeutic biomarkers, including CD4+, CD8+, CD8+ effector memory and CD8+ Naïve T-cells, naïve and unswitched memory B-cells and total CD14+ monocytes. This work could contribute to the evolution of more precise diagnostic immunological signatures for heterogeneous autoimmune rheumatic diseases such as JSLE and could facilitate better stratification of patients for optimal treatment choices and improve quality of life for patients.

## Results

### JSLE patients have a different immunological architecture to healthy controls

To investigate the potential immunological processes driving JSLE pathogenesis, in-depth immunophenotyping was performed on PBMCs from 67 patients with JSLE and 39 age-matched healthy controls (HCs) (demographic and clinical characteristics of the study cohort are given in Supplementary Table 1). Gating strategies and description of immune phenotyping panel are shown in Supplementary Figure 1 and Supplementary Table 2.

**Table 1:** Selection of important immunological features from ML analysis. 28 immune cell subsets involved in the immunopenotyping of HCs and JSLE patients assessed by flow cytometry. There are listed and ticked where: **left)** selected as top 10 most important variables in the balanced random forest (BRF) model; **middle)** selected as top 10 weighted variables in sparse partial least squares discriminant analysis (sPLS-DA) analysis; **right)** significantly associated with JSLE in logistic regression analysis. Abbreviations: Regulatory T-cells (Tregs), invariant natural killer T-cells (iNKT-cells), central memory (CM), effector memory (EM), plasmacytoid dendritic cell (PDC), Bm1 (naïve), Bm2 (mature), Bm2’ (transitional), Bm3-4 (Plasmablasts), early/late Bm5 (memory).

|  | BRF | sPLS-DA | Logistic regression |
| --- | --- | --- | --- |
| CD4+ | ✓ | ✓ | ✓ |
| CD4+ CM |  |  |  |
| CD4+ EM | ✓ |  | ✓ |
| CD4+ EMRA |  |  |  |
| CD4+ Naive |  | ✓ |  |
| CD8+ | ✓ | ✓ | ✓ |
| CD8+ CM |  | ✓ | ✓ |
| CD8+ EM | ✓ | ✓ | ✓ |
| CD8+ EMRA |  |  |  |
| CD8+ Naive | ✓ | ✓ | ✓ |
| iNKT | ✓ | ✓ | ✓ |
| Treg |  |  |  |
| Tresp |  |  |  |
| CD19+ |  |  |  |
| Bm1 | ✓ | ✓ | ✓ |
| Bm2 |  |  |  |
| Bm2 (Transitional) |  |  |  |
| Bm3-Bm4 |  |  |  |
| Early Bm5 |  |  |  |
| Late Bm5 |  |  |  |
| CD19+ Naive | ✓ |  | ✓ |
| CD19+ Switched memory |  |  |  |
| CD19+ Unswitched memory | ✓ | ✓ | ✓ |
| CD14+ | ✓ | ✓ | ✓ |
| Classical |  |  |  |
| Intermediate |  |  |  |
| non-classical |  |  |  |
| PDC's |  | ✓ | ✓ |

**Figure 1:**
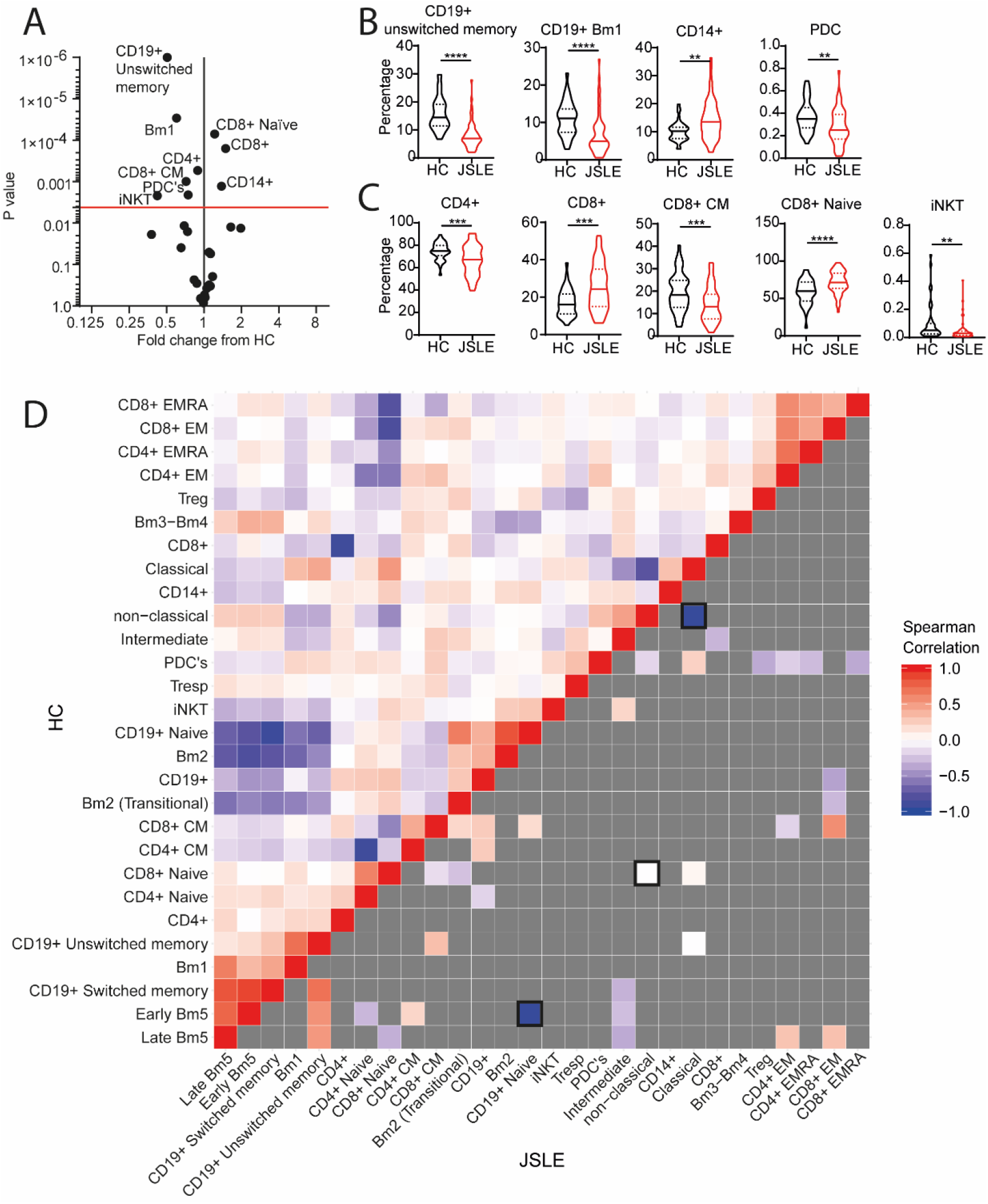
The immunological architecture is altered in JSLE: PBMC’s from HCs (n=39) and JSLE patients (n=67) were stained ex-vivo as described in Supplementary Figure 1 (gating strategy) and Supplementary Methods Table-1, to evaluate expression of 28 immune cell subsets by flow cytometry. **(A)** Volcano plot displaying data points from all 28 immune cell subsets between JSLE patients and HCs. Fold change and Log^10^ p values are displayed from unpaired t tests. Red line=adjusted p value following 1% false discovery rate adjustment for multiple comparisons. **B-C)** Violin plots displaying **B)** antigen presenting cells and **C)** T-cell subsets that were significantly different between HCs and JSLE patients by unpaired t test. Mean+SE, *=p<0.05, **=p<0.01, ***=p<0.001, ****=p<0.0001. **D)** Correlation comparison analysis was performed on immune phenotyping data described in (A). **Upper left** of the heat map: correlation between pairs of immune cell types (28 immunological variables) in HCs. Spearman correlation coefficients for each pair of cell types are represented by colour (red indicates a positive correlation coefficient; blue indicates a negative correlation coefficient). Colour concentration is proportional to the strength of the correlation (corresponding value shown on the right of the figure). **Bottom right** of the heat map: Correlation between pairs of immune cell types in JSLE patients, where a dark grey colour indicates the Spearman correlation coefficient is not signficantly different to HCs. Correlations between pairs of immune cell types which were significantly altered in JSLE patients are coloured (p<0.05) (red colour indicates a positive correlation coefficient; blue indicates a negative correlationcoefficient) and boxed (p<0.01).

Initial analysis by t-test revealed that JSLE patients had a vastly different immune cell profile compared to HCs following 1% false discovery rate (FDR) correction for multiple testing (Figure 1A-C). This included increased total and naïve CD8+ T-cells and CD14+ monocytes as well as decreased total CD4+ T-cells, CD8+ central memory (CM) T-cells, invariant natural killer T (iNKT)-cells, naïve (Bm1) and unswitched memory B-cells and plasmacytoid dendritic cells (PDC’s).

The immunological architecture of JSLE patients compared to HCs was further explored using correlation comparison analysis of the 28 immunological parameters described above. In the HC population significant correlations were observed between many different types of immune cells, most notably, significant negative correlations were identified between naïve and memory B-cell sub-populations and separately between naïve and memory T-cell populations (Figure 1D-upper triangle, and Supplementary Table 3). In JSLE, a clear global change in immunological architecture was evident compared to HCs: many of the immune cell relationships identified in HCs were inverted or exacerbated in JSLE. Most notably, an increased negative correlation between naïve and memory (Early-Bm5) B-cells, an increased positive correlation between switched and unswitched memory B-cells and an increased negative correlation between classical and non-classical monocytes. (Figure 1D-lower triangle, and Supplementary Table 3). These results suggest a comprehensive alteration of the immune system in JSLE with substantial memory lymphocyte involvement indicating dysregulation of the adaptive immune system.

### Machine learning identifies a unique immune signature associated with JSLE

To further assess the importance of the immunological composition in JSLE pathology, a balanced random forest (BRF) ML approach utilising immunophenotyping data was employed to discriminate between patients with JSLE and HCs (Figure 2A for BRF schematic and methods section for description of the approach). Each decision tree was constructed with a randomly generated subset of the original sample data, where only a random subset of the immunological parameters is considered in each split. In order to maximise the performance of the predictive model, a parameter-tuning test was performed by comparing model accuracy using different numbers of randomly selected immunological parameters in each split. BRF models with 10 immunological parameters (N_variables_=10) selected in each split gave the best overall model accuracy, and thus were employed in the following analysis. To ensure the reliable predictive performance of the model, a total of 10,000 decision trees were used for model construction.

**Figure 2:**
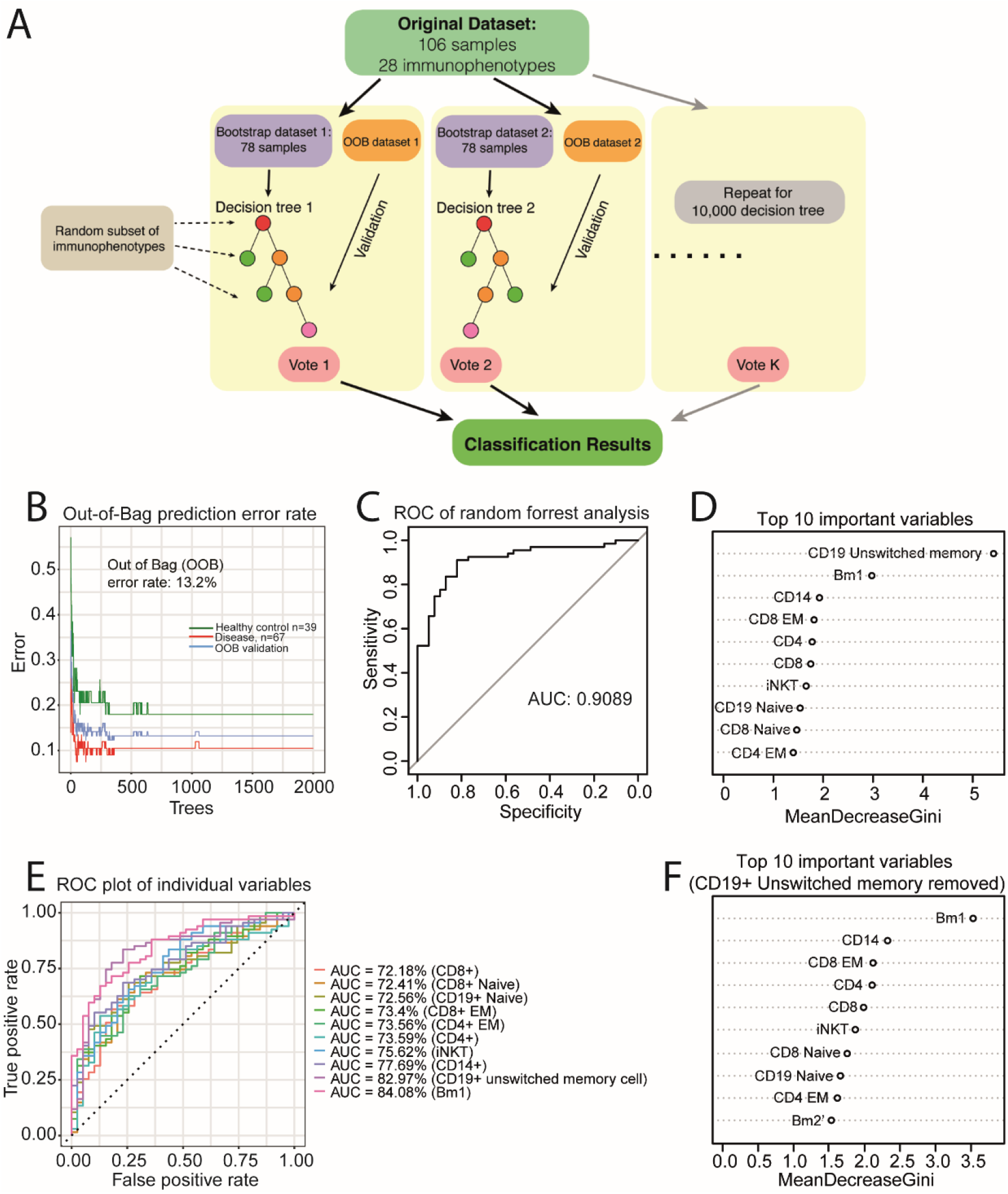
Balanced random forest machine learning using immunophenotyping data can predict accurately JSLE patients from healthy controls. **(A)** Building a predictive model using a Balanced Random Forest approach. A large number of decision trees are used (in this case n=10,000) (see methods section). Demographic variables including gender, age and ethnicity were adjusted for in the model. **(B)** Comparison of 28 different immune cell subsets in healthy donors (n=39) vs JSLE patients (n=67) using the Balanced Random Forest Model.The out of bag (OOB) error rate was 13.2%. **(C)** Reciever operator characteristic (ROC) curve analysis and 10-fold cross validation was used to validate the model providing an area under the curve (AUC) of 0.908 and an accuracy of 87.80%, meaning that the model was able to confidently predict differentiation between HCs and JSLE. **(D)** The top 10 variables contributing to the random forest model are shown. The mean decrease in Gini is a measure of each variable to the importance of the model, a higher score indicates a higher importance of the variable. **(E)** ROC showing the sensitivity and specificty of the top 9 markers identified by the model. This shows that all variables are important for the prediction. **(F)** The top 10 contributing variables in the random forest model trained on a total of 27 immunological parameters (excluding CD19+ unswitched memory cells).

After optimisation, the BRF model identified JSLE patients from HCs with a classification accuracy of 86.8% (Figure 2B). The classification error rate in the out-of-bag validation set was 10.4% and 17.9% for predicting JSLE samples and HC samples respectively. Receiver operating curve (ROC) analysis of the BRF model showed an AUC of 0.909 (Figure 2C), indicating the outstanding efficiency of the model in discriminating JSLE samples from HC samples. From this analysis the diagnostic sensitivity and specificity was 89.6% and 82.1% respectively. In addition, the classification accuracy held a steady measure of 87.8% in the 10-fold cross-validation analysis.

Thus, JSLE patients can be discriminated from HCs with high confidence using these immunological parameters and this BRF ML model.

### Important immunological parameters selected from the balanced random forest model

The top contributing feature identified by the random forest algorithm to segregate JSLE from the HC was the altered frequency of CD19+ unswitched memory cells, followed by Bm1 (naïve) B-cells and CD14+ monocytes (Figure 2D). To further evaluate the influence of individual immunological parameters, individual random forest models using each of the most important variables were performed. The AUC of these univariate random forest models ranged from 72.18% to 84.08%, with the best performance given by the Bm1-only model (AUC=0.8408), followed by the CD19+ unswitched memory B-cell-only model (AUC=0.8297) (Figure 2E). These results confirm the important role of each primary immunological parameter selected in the original random forest model.

Removing individual parameters from the random forest model was investigated; the distribution of important variables generated from the model that excluded CD19+ unswitched memory B-cells was similar to that of the original model (Figure 2F). Therefore, the individual frequency alteration of CD19+ unswitched memory B-cells and other select immunological parameters might not be the critical explanation for JSLE pathogenesis. Instead, the systemic alteration of the immune system involving multiple immune cell dysregulations might better explain the complexity and heterogeneity of the disease phenotype.

### Validation of the top immunological parameters associated with JSLE from the balanced random forest model

To further evaluate and validate the relationship between the individual immunological parameters and JSLE, logistic regression analysis was applied by modelling the probability of JSLE using the immune profiles of the HC and JSLE cohorts. Odds ratios (ORs) and 95% confidence intervals produced by the logistic regression model were used to evaluate the strength of the association between individual immunological parameters and JSLE (Figure 3A, Supplementary Table 4). In agreement with the results of the autoregulation analysis (Figure 1D), logistic regression analysis identified 12 out of 28 immune cell types as significantly associated with JSLE, substantiating the global immunological difference between JSLE and HCs. The correlation between having JSLE and the reduced frequency of CD19+ unswitched memory cells was relatively high (OR=0.71, 95%CI = (0.60, 0.82)), in accordance with previous BRF classification analysis (Figure 2D). Variables that were selected by the optimal BRF model were all confirmed to be significantly associated with JSLE by logistic regression, thus strengthening the evidence of their potential application as diagnostic biomarkers. Of note, vastly significant negative associations were observed in iNKT-cells (OR = 4.59e-06, 95%CI = 1.52e-10, 0.14) and PDCs (OR=2.03e-02, 95%CI = 6.11e-04, 0.67); for this reason, they were removed from the forest plot.

**Figure 3:**
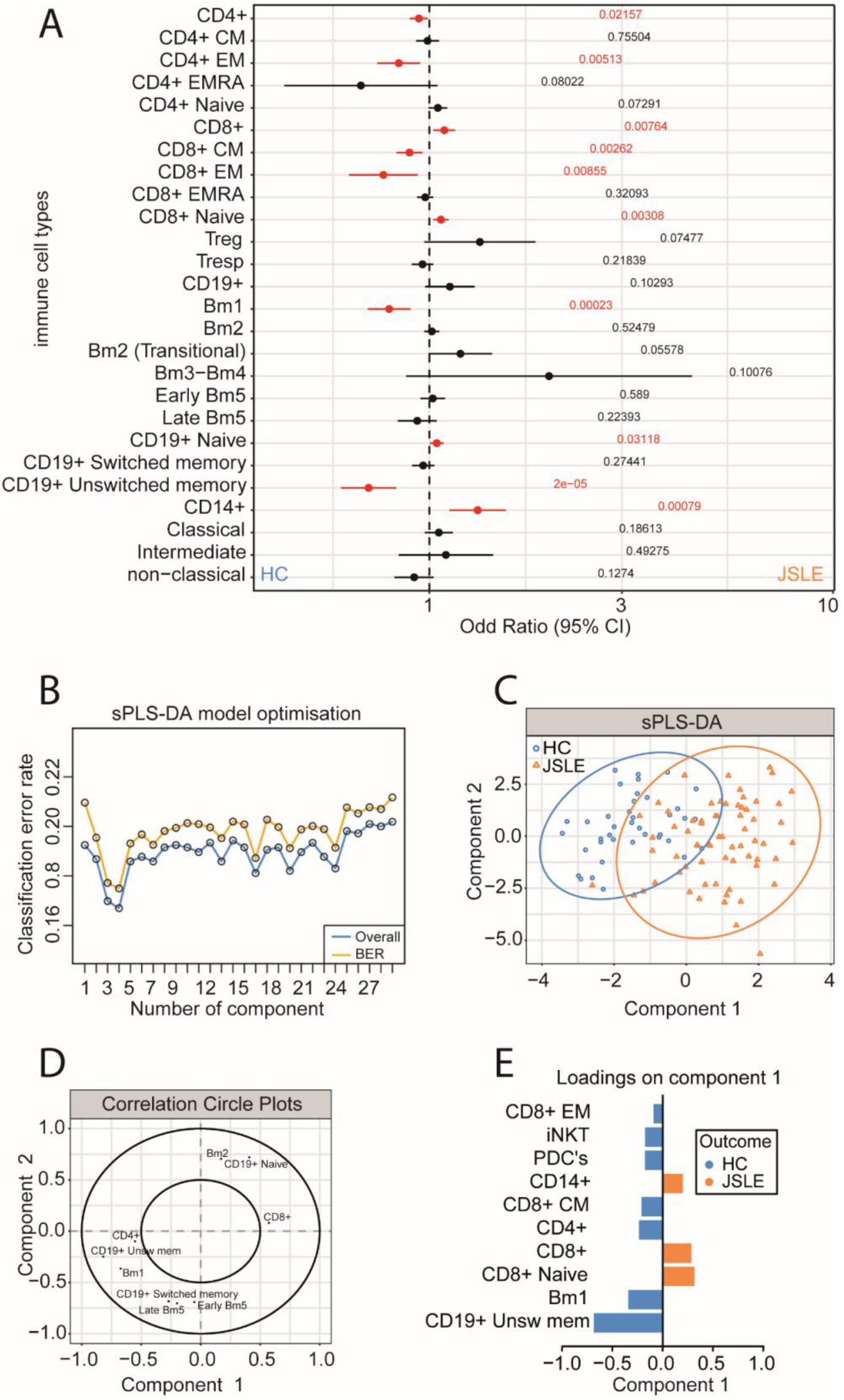
Top hits from BRF approach were validated using logistic regression analysis and sparse Partial Least Squares Discriminant Analysis (sPLS-DA). **(A)** Odds ratios (95% confidence intervals, CIs) of 28 immunological parameters were computed with logistic regression analysis. The dotted line represents the line of no effect (OR=1). 95% CIs that do not cross the dotted line (OR=1) indicate a statistically significant association between the immune cell type and JSLE (in red). Gender, ethnicity and age were adjusted in the logistic regression analysis to avoid confounding effects. For better visualization of the result, iNKT and PDC were omitted from the figure. **(B-E)** sPLS-DA analysis was perfomed. **(B)** Model optimisation. Models with different component numbers were assessed by 10-fold cross-validation ×10, using the overall error rate (blue line) and balanced error rate (BER, yellow line) to evaluate model performance. The models with four components gave the lowest overall estimation error rate (16.7%) and BER (17.5%) and were selected as optimal for sPLS-DA models. **(C)** sPLS-DA plot to validate the top hits from the predictive model. sPLS-DA is a supervised clustering method, which applies weighting to measurements which separate HCs and un-stratified JSLE patients. Ovals indicate the 80% prediction interval. Using this analysis, the weighting of each cell type in component 1 is displayed **(D)** as well as their factor loading value **(E)**. Demographic variables including gender, age and ethnicity were adjusted for in all analysis.

As a secondary validation, sparse partial least squares discriminant analysis (sPLS-DA) was performed to rank and validate the immunological variables by their distribution in JSLE and HCs. sPLS-DA is a supervised clustering machine learning approach that combines parameter selection and classification into one operation. By assessing the overall estimation error rate and balanced error rate in 10-fold cross-validation, models with four components were chosen for optimal model performance (Figure 3B). After applying the optimal immunophenotyping-aid sPLS-DA, a significant separation between JSLE and HCs was observed by plotting principal component (PC) 2 against PC 1 (Figure 3C), indicating a good prediction ability for the model. Similar to the BRF analysis, a subset of discriminating immune cell types were selected and ranked by discriminating capability (Figure 3D-E). Unsurprisingly, the highest weighted immunological parameter was CD19+ unswitched memory cell (weight = -0.69), followed by Bm1 at approximately half of the weight (-0.34). The top 10 discriminating parameters selected from sPLS-DA were all reported as significantly associated with JSLE and matched the most important parameters from the BRF model, with the exception of CD8+ CM cells and PDCs. Thus, a distinct immune signature was identified and validated that could be applied diagnostically to identify JSLE patients for faster targeted immunotherapy (Table 1).

### Patients can be stratified based on the top hits from the ML model

To assess whether the immunological signature identified could be used to further stratify JSLE patients, K-mean clustering, an unsupervised ML algorithm was used. For this analysis, immunological parameters fulfilling the following criteria were selected: 1) selected as top 10 important variables in balanced random forest model; 2) selected as top 10 weighting variables in sPLS-DA analysis; 3) significantly associated with JSLE in logistic regression analysis. After screening, eight immune cell types stood out from the 28 immunological parameters: CD4+, CD8+, CD8 EM and CD8+ Naïve T-cells, Bm1 and unswitched memory B-cells and total CD14+ monocytes (Table 1). K-mean clustering was performed to partition the original JSLE patients into four groups (Group(G)1, n=10; G2, n=21; G3, n=21; G4, n=15) based on the top-weighted immunophenotyping variables, with distinct immune profiles (Figure 4A and B).

**Figure 4:**
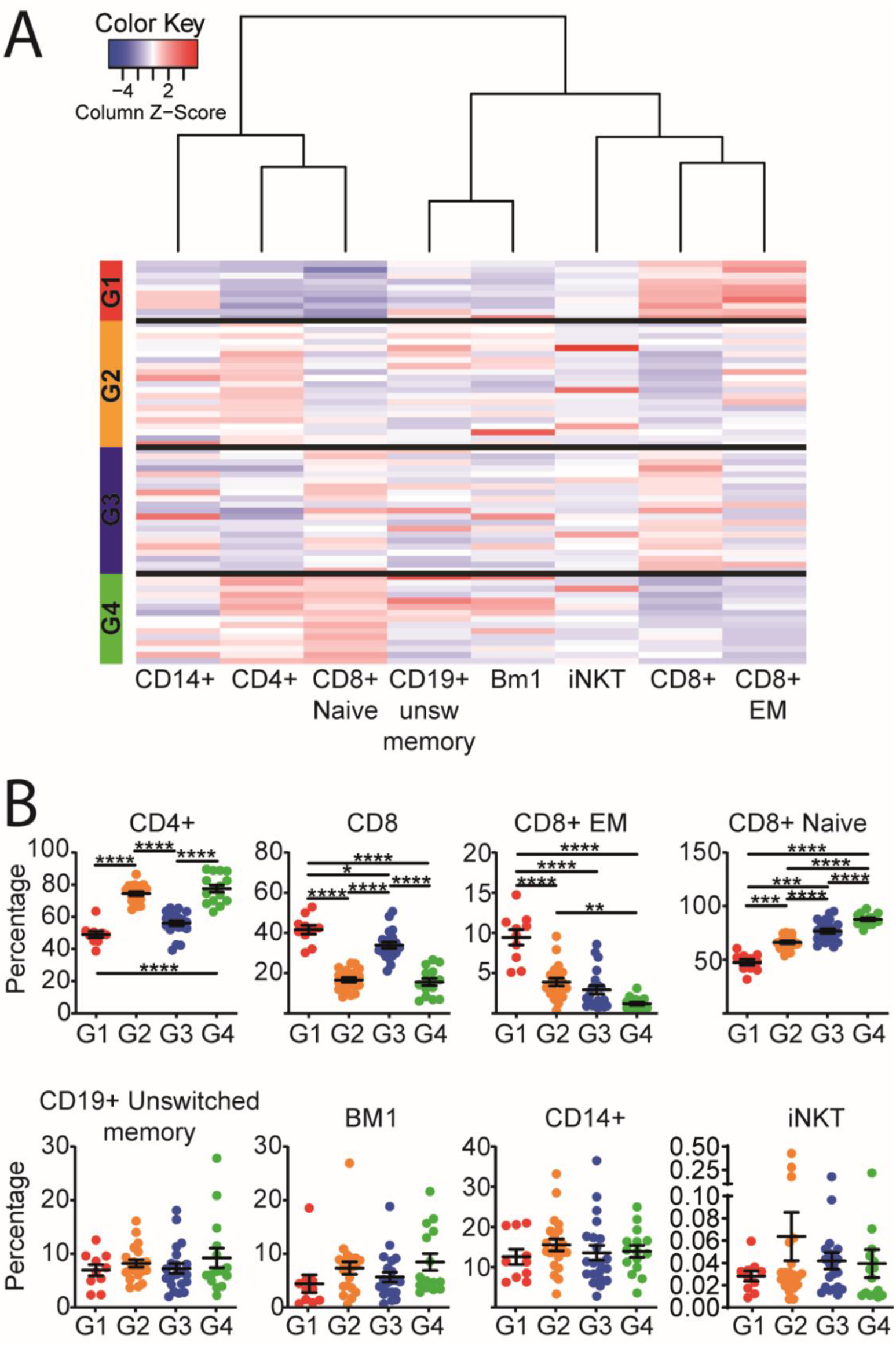
Patient clustering by top-weighted immunological parameters in JSLE compared to HCs. **(A)** JSLE Patients were stratified using k-means clustering including the using immunophenotypes of the selected top-weighted immunological parameters. The immunophenotypes are plotted as a heatmap, each row representing an individual JSLE sample, which are clustered by groups. The immunophenotype is standardised within each column by z-score. The colour key shows the corresponding colour for z-score (from -5 to 5) for each column, representing the relationship to the mean of the group; red colour indicates a relatively high frequency of the immune cell type, and blue colour indicates a relatively low frequency of the immune cell type. Four groups of patients were recognised with distinct immune cell profiles. Group 1 (n=10, red); Group 2 (n=21, orange); Group 3 (n=21,blue); Group 4 (n=15, green). **B)** Scatter dot plots displaying top-weighted immunological parameters between the k-means clustered groups (coloured appropriately). Mean+SE, one-way ANOVA, Tukey’s multiple comparisons test, *=p<0.05, **=p<0.01, ***=p<0.001, ****=p<0.0001.

Clear patterns among T cell subsets were observed from the patient grouping, with significant differences in CD4+, CD8+, CD8+ naïve, and CD8+ EM T cell subset frequencies (Figure 4B). JSLE samples in group (G)1 and G3 share relatively high CD8+ T-cells and relatively low CD4+ T-cells, whereas the opposite phenotype was observed in G2 and G4. G1 also had relatively high EM but low naïve CD8+ T-cells. Interestingly, no significant differences were identified between the groups for unswitched memory and Bm1 B-cells, CD14+ monocytes or iNKT cells. In addition, relative comparison revealed no demographic or clinical differences between any groups with the exception of complement component 3 (C3) which was significantly reduced in G1 and G3 (Supplementary Table 5). Thus, despite the identification of a strong predictive immune signature associated with JSLE, patient heterogeneity in JSLE could still be important for considering targeted and tailored treatment.

### Immune cell subsets associated with JSLE are driven by different clinical features

Finally, in order to explore the systemic association between immunological parameters and serologic clinical features a network analysis was performed. Clinical features including the measurement of disease activity (SLE disease activity index, SLEDAI), anti-double stranded (ds)DNA antibodies, complement levels, serum lipids, and other JSLE-associated biomarkers, were selected to examine the relationship between immune cell frequencies and JSLE disease features. From the overall correlation network, extensive immune correlations across clinical features were observed (Figure 5, Supplementary Table 6). Consistent with the previous top-weighted immunological parameters (Table 1), frequent connections involved total CD4+ and CD8+ T-cells, naïve CD8 T-cells and Bm1 and unswitched memory B-cells, whereas iNKT-cells only correlated positively with body mass index (BMI) and C3.

**Figure 5:**
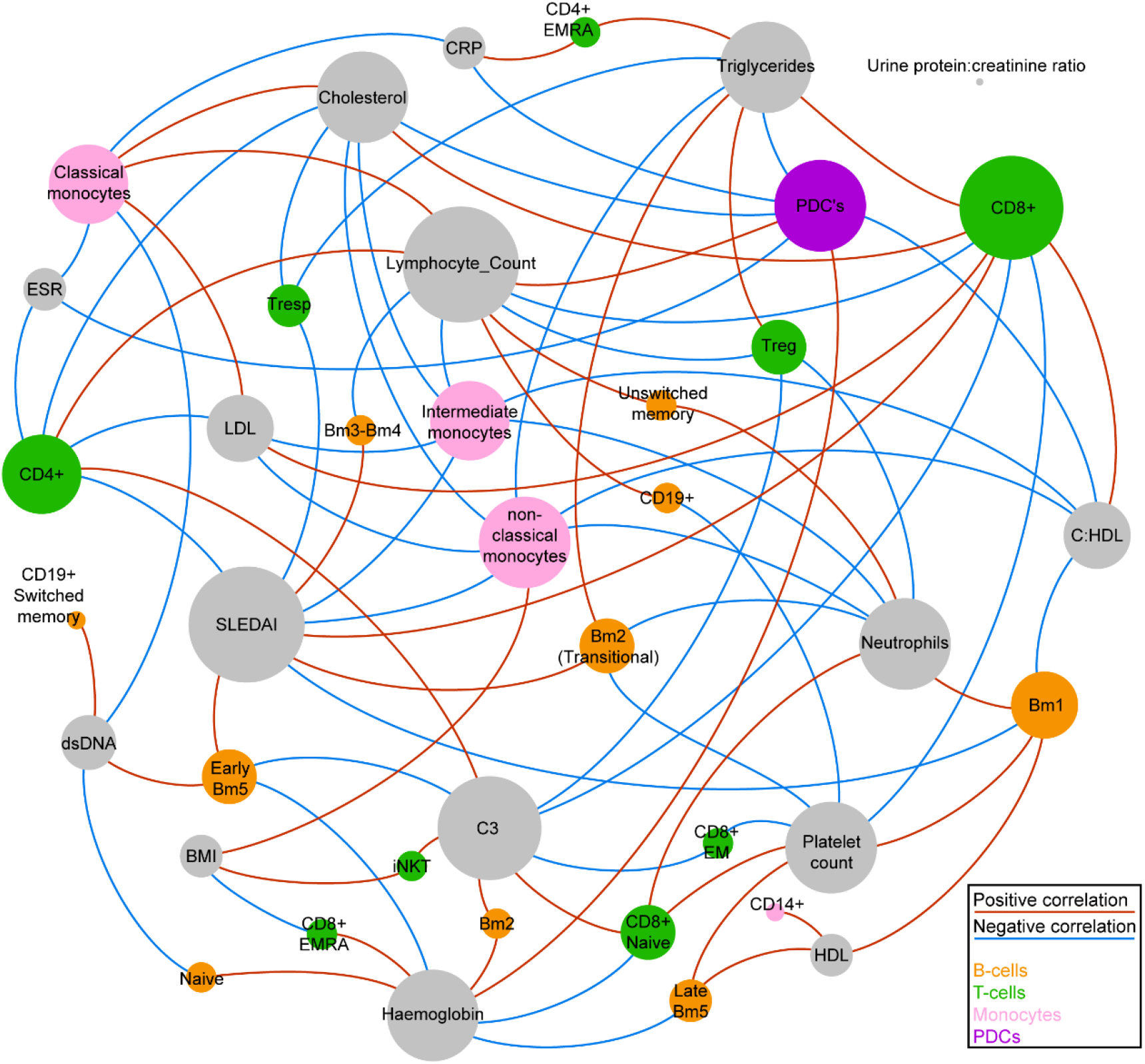
Network analysis identifies associations between immunological and clinical features JSLE. Correlations between immune cell frequency measures and JSLE clinical characteristics. Pearson correlation coefficients based on univariate logistic regression are represented as connecting lines between the clinical characteristic nodes and immune cell frequency nodes. Only correlations with an absolute r value of 0.2 and above are shown. Immune cell subsets with no correlation (r) values above 0.2 are not shown. P values and r values are displayed in Supplementary Table 4. Size of the circles (nodes) are proportional to the total number of connections with other nodes. Red line colour indicates a positive correlation and blue lines indicate a negative correlation. Node colour was grouped according to immune cell type (T-cells: green, B-cells: orange, monocytes: pink, PDC’s: purple) and clinical characteristic (grey). The graph was generated using the Force Atlas layout in Gephi 0.9.2.

Disease activity, (SLEDAI) correlated negatively with CD4+ T-cells, naïve B-cells and intermediate and non-classical monocytes and positively with CD8+ T-cells, transitional and mature B-cell subsets. Anti-dsDNA antibody measures correlated with early Bm5 and switched memory B-cells with no correlation with T-cell subsets. C3 also had many significant correlations, including a negative correlation with total and EM CD8+ T-cells, supporting patterns associated with the k-mean clustered groups (Figure 4 and Supplementary Table 5).

These results suggest the potential interaction between clinical features and disease-related immune dysregulation, which could help explain the multifactorial, heterogeneous and systemic nature of the disease.

## Discussion

While the immunological differences between HCs and patients with adult SLE have been well established in previous studies [3, 22], the immune phenotype of patients with JSLE is less well described. Defects in immune cell subtypes in JSLE may contribute to an increased disease severity and pathogenesis via different immunological pathways. Due to the complexity of the pathogenesis and the heterogeneous nature of JSLE clinical manifestations, imprecise diagnosis and poor treatment efficacy remain two major obstacles that drastically affect the outcome of patient with JSLE [2]. Here the immune profiles of 28 immune cell subsets in JSLE patients were compared to HCs to explore the immunological signature of the disease and assess potential drivers of JSLE immunopathogenesis. This analysis identified i) a global shift of the immunological architecture away from HCs in JSLE patients confirmed by correlation comparison analysis; ii) JSLE patients were distinguished from HCs using an optimal BRF model, indicating the potential immunologically-directed diagnostic power of ML; iii) Patients were clustered into four distinct groups based on the top hits from the ML model and iv) specific clinical features were differentially associated with immune cell subsets in JSLE. Together, these findings contribute to JSLE diagnostic research, help identify meaningful therapeutic targets for JSLE treatment and uncover the underlying pathogenic mechanisms of JSLE.

Precise and early diagnosis of JSLE is particularly important because morbidity and mortality can be significantly controlled with suitable treatment [23]. A recent study showed that in the UK, JSLE diagnosis based on the onset of symptoms is accomplished in a median time of two months, but there was a significant variation in diagnosis time (interquartile range 1–6 months) due to the highly heterogeneous clinical manifestation of the disease [24, 25]. In clinical practice, 38% of SLE patients are diagnosed with at least one other co-occurring autoimmune disease [26]. Therefore, symptoms common in typical SLE patients such as fatigue, hair loss and lymphadenopathy are normally indeterminate for JSLE diagnosis as they also present in multiple other diseases. To accurately diagnose JSLE, a variety of clinical features and laboratory markers are necessary in order to distinguish JSLE from other systemic autoimmune conditions, such as juvenile idiopathic arthritis and juvenile dermatomyositis, and requires expertise across multiple fields.

An initial SLE diagnosis is highly dependent on disease presentation without comprehensive laboratory testing [27], which makes the diagnosis of patients with inconspicuous symptoms particularly challenging. Considering the importance of immunophenotyping assessments and autoantibody tests, it is becoming crucial to include comprehensive peripheral blood tests in first-and second-line diagnostics [28]. The limited number of selected immune parameters makes the immune-based JSLE diagnostic assay economically feasible [29, 30]. ML models with substantial predictive accuracy can assist clinicians with complicated diagnostic decision making, though extensive study is necessary to construct accurate models for specific population groups. The autonomy of ML is attributable to their exceptional learning ability, which allows for continuous self-improvement without having step-by-step instructions.

Using ML approaches here, 8 out of 28 of the immune subtypes were confirmed as significantly associated with JSLE, and they contribute substantially to the classification models. In principle, these select immunological features can be further assessed as valuable biomarkers and designed as a potential diagnostic tool. In addition, the high percentage (steady measure of 87.8%) obtained from the 10-fold cross validation suggests a consistently reliable model performance without overfitting issues when applied in practice. This tool could enhance current diagnostic routines by providing a more in-depth view of the patient’s immunological state.

The development and use of biologic therapies have been increasingly promising because they can target specific immunopathological agents and diminish SLE disease progression [31]. However, the efficacy of each biologic drug is unsatisfactory due to the heterogeneous immunopathogenesis [23]. For instance, the complete response (CR) rate of rituximab assessed by the British Isles Lupus Assessment Group (BILAG) was only 46.7% for SLE [32]. The importance of the treat-to-target approach for SLE patients is addressed by van Vollenhoven and colleagues [33], as the appropriate target can be identified and resolved with specific therapy for each patient. In addition, biological drugs have become increasingly popular in routine management of patients with severe JSLE, but none have been approved for use in paediatric patients [23]. Despite the strong predictive immune signature developed from our ML method, JSLE patients could still be clustered into four distinct groups based on the expression of the top immune cell subsets from the ML model. Significant differences between the groups were only observed for T-cell subsets which may be associated with C3 levels; less heterogeneity was observed in the frequency of unswitched memory and Bm1 B-cells and CD14+ monocytes, which may explain the increased predictive power of these subsets in the BRF model. Therefore, using immunological biomarkers to unravel the underlying disease mechanism may allow for a more personalised approach, improving clinical trial design, approval rate and treatment efficacy of JSLE patients.

This project has a number of potential limitations. While ML approaches are well-known for their robustness and their capability to self-learn, many ML algorithms function like an incomprehensible “black box” that produces reasonable prediction results without giving complementary justification [34]. Since this work was designed to be heavily based on ML, the lack of a rational biological explanation from the results is an inevitable problem. Therefore, to uncover the immunopathogenic significance from selected immune features, a longitudinal study of JSLE cohorts is critically important for validating the association between immune cell subsets and disease.

Patients with JSLE often have overlapping clinical manifestations with other autoimmune diseases, including Sjögren’s syndrome and Rheumatoid arthritis (RA) [35]. Due to the limited cohort recruited for this study, patients with other autoimmune diseases were not included in the analysis. Therefore, the immunological differences between JSLE and other immune diseases were not assessed. Additionally, ML models were only trained with JSLE and HC samples. As a result, the performance of classification models is limited when applied to the overall population where other confounding diseases are present. Separating patients with autoimmune diseases that show extensive overlapping of symptoms will be particularly meaningful in precision diagnosis.

In conclusion, this study investigated the ability of immunophenotyping to stratify patients with JSLE and demonstrated the potential of immune-based ML to develop precision diagnosis and personalised treatment. By applying immune-based ML approaches, a BRF algorithm discriminated JSLE patients from HCs with an accuracy of ∼91%. Future multi-omics association studies have the potential to investigate the correlation between the essential immunological parameters and clinical manifestations, extracting the full value of the immune signatures in clinical practice.

## Materials and Methods

### Patients and control samples

Patient blood was collected from JSLE patients (n=67) attending a young adult or adolescent rheumatology clinic at University College London Hospital (UCLH) or Great Ormond Street Hospital (GOSH) respectively. Teenage and young adult healthy control (HC) blood (n=39) was collected at the Rayne building, UCL, from volunteers taking part in educational events such as young scientist days. Informed consent was acquired from both patients and healthy controls under the ethical approval reference: REC11/LO/0330. All information was stored as anonymised data. Detailed clinical characteristics and disease features were recorded from patient files and questionnaires (Supplementary Table 1). Disease activity was calculated via the SLE Disease Activity Index (SLEDAI). A score of 4 or more was used to indicate active disease [36].

### Flow cytometry

#### Surface staining

Multiparameter flow cytometry was used to immunophenotype JSLE and HC PBMCs stained with fixable blue dead cell stain (ThermoFisher) and a T-cell and antigen presenting cell (APC) antibody panel to measure cell marker expression (28 immune cell subsets, Supplementary Figure 1 for gating strategy, Supplementary Methods Table 1 for list of antibodies used). PBMCs were stained at a density of 1×10^6 cells per well (96 well round bottom plate). This was followed by subsequent washes and fixation in 2% PFA before running on a flow cytometer.

#### Data acquirement and analysis

Data was acquired using a BD LSRFORTESSA X-20 flow cytometer (1×10^6-2×10^6 cells per sample) and analysis was carried out using FlowJo Single Cell Analysis Software (TreeStar). Application settings were created to begin with and Cytometer Setup and Tracking (CS&T) (BD) beads were run on the flow cytometer before each session to assess cytometer performance. Application settings were applied to panel templates each time prior to compensation to ensure that all immunophenotyping data was comparable over time. Gating strategies were kept consistent throughout and are shown in Supplementary Figure 1.

### Statistical analysis

The clinical features and immune profile data of 67 JSLE patients and 39 HCs were collated and stored in Microsoft Excel. Individual identifications were replaced with randomly assigned numbers to ensure confidentiality. General immunophenotype data compared between HCs and JSLE patients or across JSLE stratified groups was performed by unpaired t-test using GraphPad Prism 8 software and plotted as violin plots, scatter dot plots or as a volcano plot where data was corrected for multiple testing by two-stage linear step-up procedure of Benjamini, Krieger and Yekutieli (false discovery rate (FDR) of 1%). All other statistical analyses were performed in R version 3.5.2 [37] and demographic variables including gender, age and ethnicity were adjusted for in all ML analyses. The software/packages used for processing any analysis are listed in Supplementary Methods Table 2 and are detailed below.

#### Correlation Comparison analysis

Correlation comparison analysis was performed to investigate the difference in immunological architecture between the HC and the JSLE patients. Spearman correlation tests between pairs of immune cell types in both the HC and the JSLE patients were performed using R version 3.5.2 [37]. The significance for the difference in corresponding correlation between the HC and the JSLE patients was calculated using the cocor package in R (*cocor*.*indep*.*groups* function) [38]. Spearman correlation coefficients for pairs of immune cell types were plotted in a heat map using the heatmap.2 function from gplots package in R (Supplementary Methods Table 2).

#### Balanced random forest (BRF)

To stratify JSLE patients from the HC using immunophenotyping data, the balanced random forest (BRF) approach was used with the randomForest package in R (Supplementary Methods Table 2). Decision trees were built using a bootstrap dataset consisting of randomly selected samples from the original dataset (n = 106), allowing the same sample to be selected more than once. As the original sample set had an unbalanced HC:JSLE (39:67) ratio, the balanced method was applied in the bootstrap dataset construction. The bootstrap dataset was first selected from the minority class (HC, n = 39) whilst randomly drawing the exact number (n = 39) from the majority class (JSLE). The balanced bootstrap dataset (n = 78) was then used for model training. After creating the bootstrap dataset, only a random subset of immunological variables was considered at each split of the decision tree. Every decision tree was built by constructing a new bootstrap dataset and considering a newly selected subset of variables at each step. A total 10,000 decision trees were used for the BRF model construction, allowing the output to be stabilised. The classification output of the model was provided by aggregating the predictions of every decision tree and making the final prediction. Samples that were not included in the bootstrap dataset were termed the Out-of-Bag (OOB) dataset and were used to validate the model performance.

In addition, model optimisation testing was performed to determine the exact number of immunological variables (N_variable_) included in each subset for building the decision tree. By comparing the accuracy of the BRF model with different Nvariable settings (N_variable_ ranging from 1 to 28), the model with the overall lowest classification error rate was selected as the optimal model and applied in further analysis. For model performance evaluation, the ROC plot and the AUC of each model was computed with the pROC package in R (Supplementary Methods Table 2). To assess the BRF model performance in practice, 10-fold cross validation was applied with the caret package in R (Supplementary Methods Table 2).

#### Sparse partial least squares discriminant analysis (sPLS-DA)

This supervised machine learning approach sPLS-DA was operated using the mixOmics package (Supplementary Methods Table 2) in R. Ten-fold cross-validation with 50 repetitions was applied to prevent model overfitting. Model optimisation was applied to select the number of components included in the sPLS-DA model. By comparing the overall estimation error rate and the balanced error rate (BER) from 10-fold cross-validation, the model with the best discriminatory performance was selected as the optimal model and was used for further analysis. The separation of JSLE and HC samples was presented by projecting the samples into the subspace constructed of component 1 and component 2. The prediction interval of the model was calculated from the 95% confidence ellipses for the HC group and the JSLE group. The top 10 weighted immunological parameters were selected and presented by variable loading plots.

#### Logistic regression for association analysis

The association between the immunophenotypes of 28 parameters and JSLE was assessed by logistic regression analysis adjusted for age, sex and ethnicity. For each measurement, the odds ratio (OR) and the 95% confidence interval (CI) were determined. The p-value for each association was calculated in the logistic regression analysis. Forest plots produced with the ggplot2 package (Supplementary Methods Table 2) in R were used to present the logistic regression analysis results, with significant associations highlighted in blue (p < 0.05).

#### K-means clustering analysis

To apply the identified biomarkers in JSLE patient stratification, k-means clustering analysis was performed with the stats package in r. The k-means clustering algorithm repositions the specific amount of cluster centroids around the JSLE samples (n=67) until the most convergent grouping appears. The number of groupings in k-means clustering is determined by the elbow method (Supplementary Methods Figure 1). The immunophenotypes of the selected top hits parameter were standardised and laid out as a heatmap, with columns representing selected immune cell types and rows representing individual JSLE samples.

#### Network analysis

To visualise the systemic connection between immune cell type and clinical features, network analysis was performed with Force Atlas layout in Gephi (Supplementary Methods Table 2). 16 clinical features, namely SLEDAI score, cholesterol, high-density lipoprotein (HDL), C:HDL, low-density lipoprotein (LDL), triglycerides, lymphocyte count, complement component 3 (C3), C-reactive protein (CRP), double-stranded DNA (dsDNA), erythrocyte sedimentation rate (ESR), haemoglobin, platelet count, urine protein:creatinine ratio, neutrophil count and body mass index (BMI) were applied in the network analysis, covering essential measurement of disease activity, lipid levels, inflammation and autoantibody levels. Pearson correlation coefficients for each association were calculated in R. Only correlations with an absolute value of ≥0.2 are shown in the graph.

## Data Availability

All relevant data is available within the manuscript

## Contributors

Design of research study; ECJ, CC, ITP, PD. Acquiring data; GAR, AR, HP, CW. Recruiting patients; HP, CC, YI, CW, DAI. Analyzing data; GAR, JP, PD, LC. Writing the manuscript; GAR, JP, ECJ. Review of the manuscript; DAI, CC, ITP, PD. All authors approved the final version.

## Ethics approval

This study was approved by the London-Harrow research Ethics Committee,11/LO/0330

